# Explainable machine learning models to understand determinants of COVID-19 mortality in the United States

**DOI:** 10.1101/2020.05.23.20110189

**Authors:** Piyush Mathur, Tavpritesh Sethi, Anya Mathur, Kamal Maheshwari, Jacek B Cywinski, Ashish K Khanna, Simran Dua, Frank Papay

## Abstract

**Background:** COVID-19 is now one of the leading causes of mortality amongst adults in the United States for the year 2020. Multiple epidemiological models have been built, often based on limited data, to understand the spread and impact of the pandemic. However, many geographic and local factors may have played an important role in higher morbidity and mortality in certain populations.

**Objective:** The goal of this study was to develop machine learning models to understand the relative association of socioeconomic, demographic, travel, and health care characteristics of different states across the United States and COVID-19 mortality.

**Methods:** Using multiple public data sets, 24 variables linked to COVID-19 disease were chosen to build the models. Two independent machine learning models using CatBoost regression and random forest were developed. SHAP feature importance and a Boruta algorithm were used to elucidate the relative importance of features on COVID-19 mortality in the United States.

**Results:** Feature importances from both the categorical models, i.e., CatBoost and random forest consistently showed that a high population density, number of nursing homes, number of nursing home beds and foreign travel were strongest predictors of COVID-19 mortality. Percentage of African American amongst the population was also found to be of high importance in prediction of COVID-19 mortality whereas racial majority (primarily, Caucasian) was not. Both models fitted the data well with a training R^2^ of 0.99 and 0.88 respectively. The effect of median age,median income, climate and disease mitigation measures on COVID-19 related mortality remained unclear.

**Conclusions:** COVID-19 policy making will need to take population density, pre-existing medical care and state travel policies into account. Our models identified and quantified the relative importance of each of these for mortality predictions using machine learning.

## Introduction

COVID-19 pandemic shares some similarities with previous influenza pandemic but appears to be more infectious with a longer incubation period, higher percentage of patients requiring hospital care and higher mortality. The potential for asymptomatic spread combined with lack of vaccine and lack of effective treatment has resulted in devastating human and economic consequences globally.

The COVID-19 pandemic exhibits an uneven geographic spread which leads to a locational mismatch of testing, mitigation measures and allocation of healthcare resources (human, equipment, and infrastructure).^1^ Many local, national and global organization are collecting enormous amounts of data to make critical logistic decision but interpreting data and acting upon it in near-real time remains a challenge. According to the Center for Disease Control(CDC) as of April 7, across the United States(US) cumulative incidence of COVID-19 ranged from 20.6 to 915.3 cases per 100,000 and 7-day increases in incidence varied considerably from 8.3 to 418.0. ^2^

It is unclear why the number of COVID-19 cases and related deaths are higher in certain regions of the country but needs to be better understood to facilitate resource allocation.^3^ In the absence of effective treatment, understanding and predicting the spread of COVID-19 is unquestionably valuable for public health and hospital authorities to plan for and manage the pandemic.^4^ Modern machine learning techniques can identify useful patterns from diverse data sources in real-time and help understand the difference in disease spread.^5^ The features or determinants affecting COVID-19 mortality should be easily explainable to trust the predictive algorithms output and to build confidence in decision making.

While there have been many models developed to predict mortality, the authors sought to develop machine learning prediction models that provide an estimate of the relative association of socioeconomic, demographic, travel, and health care characteristics of COVID-19 disease mortality among states in the United States.

## Methods

State-wise data for the 50 states and the District of Columbia in the United States was collected for all the features predicting COVID-19 mortality and for deriving feature importance (eTable 1 in the Supplement).^6^

We excluded the territories of the United States due to limitations with reporting of data. Key feature categories include demographic characteristics of the population, pre-existing healthcare utilization, travel, weather, socioeconomic variables, racial distribution and timing of disease mitigation measures (Table 1). These features have independently been postulated to have correlation with COVID-19 prevalence and its associated mortality.

**Table 1.** **Description of the determinants of COVID-19 mortality in the United States**.

| **Feature name** | **Range (Min-Max)** | **Median** | **Interquartile Range** |
| --- | --- | --- | --- |
| Hospital beds(per 1000 population) | 1.6 - 4.8 | 2.5 | 2 - 3 |
| Doctors (per 100,000 population) | 134 - 449.5 | 263 | 229 - 298 |
| Number of nursing homes | 18 - 1217 | 226 | 84 - 383 |
| Total number of nursing home beds | 693 - 136000 | 24558 | 6949 - 38621 |
| Average household income(\$) | 41659 - 69529 | 51516 | 48267 - 60713 |
| Total population | 578759 - 39512223 | 4467673 | 1789606 - 7446805 |
| Population density (per square mile) | 1 - 11815 | 108 | 51 - 224 |
| Timing of educational facilities closure | 03/16/2020 - 04/02/2020 | NA | NA |
| Timing of stay at home order(Month) | None/March/April | NA | NA |
| Direct flights from Wuhan* | Yes/No | NA | NA |
| Direct flights from China ** | Yes/No | NA | NA |
| Direct flights from Europe*** | Yes/No | NA | NA |
| Average temperature in Jan(degrees F) | -6 - 75 | 35 | 27 - 42 |
| Average temperature in Feb(degrees F) | 1 - 75 | 36 | 26 - 42 |
| Average temperature in Mar(degrees F) | 12 - 75 | 45 | 37 - 51 |
| Absolute humidity in Jan(gram/m<sup>3</sup>) | 2 - 15 | 5 | 4 - 7 |
| Absolute humidity in Feb(gram/m<sup>3</sup>) | 2 - 13 | 5 | 3 - 6 |
| Absolute humidity in Mar(gram/m<sup>3</sup>) | 3 - 15 | 6 | 4 - 8.5 |
| Median age(years) | 31 - 45 | 39 | 37 - 40 |
| Percent of male population(%) | 47 - 51 | 49 | 49 - 50 |
| Percent of african americans(%) | 0 - 45 | 7 | 3 - 14.5 |
| Racial majority**** | Caucasian:<br>Yes/No | NA | NA |
| COVID-19 positive | 365 - 312977 | 7441 | 3442 - 21904 |
| Death | 7 - 18909 | 288 | 77 - 1097 |

NA, not applicable
\*Direct flight from Wuhan to a state in the US: 2 states(New York and California).
\*\*Direct flight from China to a state in the US: 10 states in US have direct flights from China
\*\*\*Direct flight from Europe:17 states in the US have direct flights from China.
\*\*\*\* Racial majority:Caucasian is the majority race in 46 states.(California and New Mexico,District of Columbia,Hawaii and New Mexico have Hispanic,African American,Asian race as majority, respectively)

Two machine learning models, CatBoost regression and random forest were trained independently to predict mortality in states.^7,8^ Data in both the models was partitioned into a training (80%) and test (20%) set.

CatBoost is a machine learning algorithm for gradient boosting on decision trees.^8^ This recently developed method is especially advantageous in handling complex data with categorical features avoiding the need for numerical conversion, limited need for parameter tuning and thus has high computational efficiency.^8^ Regression is a commonly used method to analyze multi-variable data and make predictions.

Random forest is a type of ensemble method which make predictions by combining the predictions of several decision trees, each of which is trained in isolation.^7^ Random forest regression is commonly used to build prediction models.It works using a combination of tree predictors such that each tree depends on the values of a random vector sampled independently and with the same distribution for all trees in the forest.^7^

Accuracy of models was assessed by R^2^ score. A commonly used measure to assess variation in the dependent variable that is accounted for by the independent variable.^9^ An R^2^ of 1.0 indicates that the data perfectly fit the linear model. Any R^2^ value less than 1.0 indicates that at least some variability in the data cannot be accounted for by the model.

Feature importance is part of a commonly used method in machine learning called feature engineering.It is used to identify the relative association of features to the model output.In our study,we used feature importance to identify the same while our model was developed to predict COVID-19 mortality in the US.Importance of the features for prediction of mortality is calculated via two machine learning algorithms - SHAP (SHapley Additive exPlanations) calculated upon CatBoost model and Boruta, a random forest based method trained with 10,000 trees for calculating statistical significance.^10,11^

SHAP is a computational algorithm based on game theory solution.^11^ It applies to the entire class of features in an additive manner and proposes SHAP values as a measure of feature importance. SHAP has optimized functions for interpreting tree-based models and a model agnostic explainer function.It affords improved computational efficiency, consistency with human intuition and better explanation of features compared to prior such algorithms.^11^

We employed a machine-learning approach for variable selection, the Boruta algorithm, which learns the dependencies in the data from the data itself. Boruta, being a random forest-based feature selection wrapper is amenable to a mixture of categorical and continuous predictors in the data.^10^ Certain desirable characteristics such as invariance to monotonic transformations, good performance in nonlinear datasets and the capability to auto-correct for dependencies between the variables makes it particularly useful for complex datasets with interactions. Also Boruta estimates statistical significance of the importance values that it provides by comparing resampled importance estimates and their comparison to a randomized *shadow data*.^10^ Only the features with Z-score statistically higher than the maximum achieved distribution for the shadow data (shown in green in Fig 1) were considered important.

**Figure 1.**
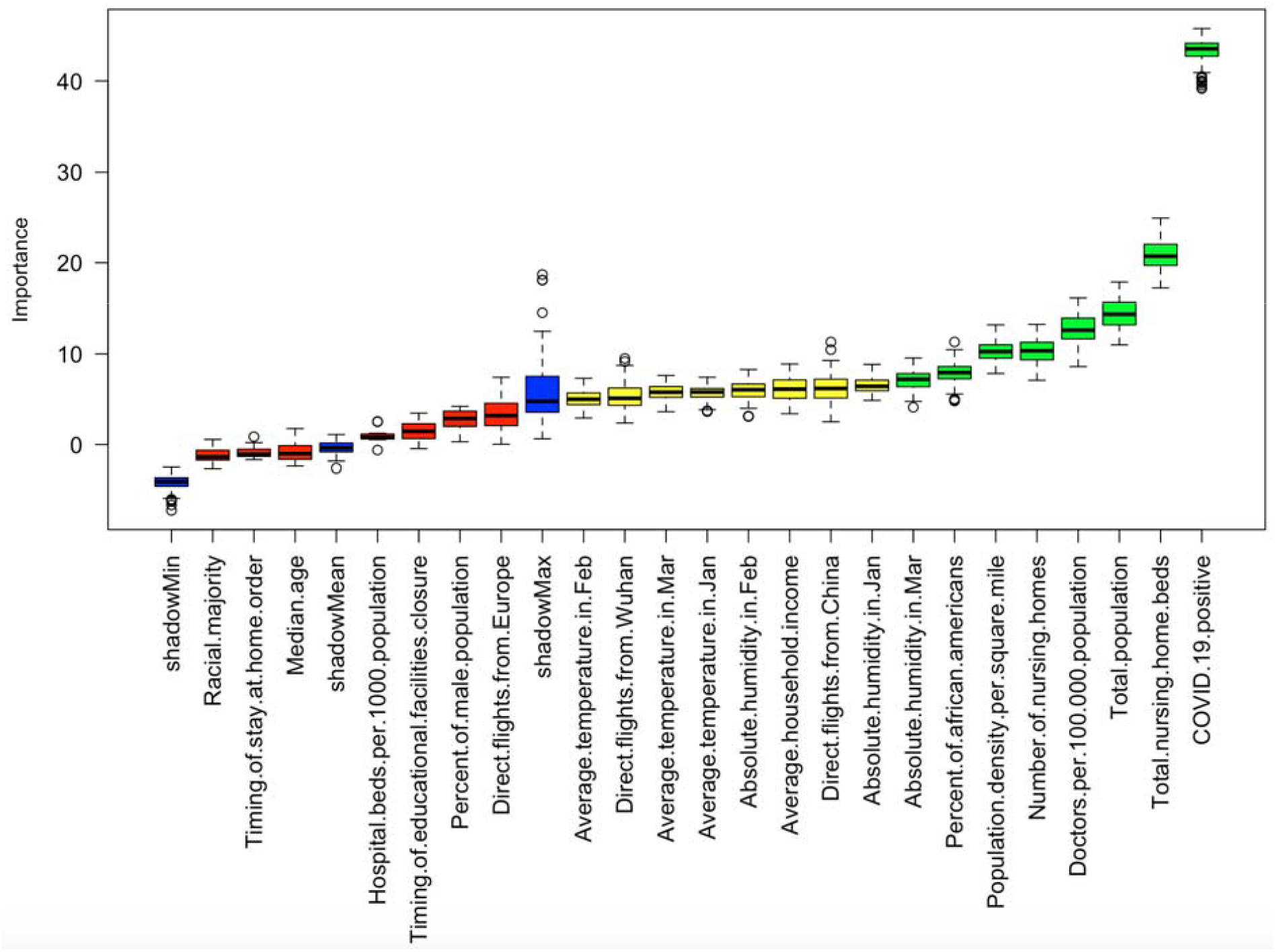
Statistical significance of feature importance computed through the Boruta algorithm for COVID-19 deaths in the US.^10^. Box plot distribution of importance, highlights features (green, yellow) whose median importance is significantly higher than the shadow importance (blue).

## Results

Results are based on 60,604 total COVID-19 deaths in the US, as of April 30, 2020. Actual number of deaths ranged widely from 7 (Wyoming) to 18,909 (New York).CatBoost regression model obtained an R^2^ score of 0.99 on the training data set and 0.50 on the test set. Random Forest model obtained an R^2^ score of 0.88 on the training data set and 0.39 on the test set.

Nine out of twenty-three variables were significantly higher than the maximum variable importance achieved by the shadow dataset in Boruta regression (Figure 1).

Both models showed the high feature importance for pre-existing high healthcare utilization reflective in the number of nursing home beds, number of nursing homes and doctors per 100,000 population(Figure 1 & 2 a, b). Overall population characteristics such as total population and population density also correlated positively with the number of deaths.Notably, both models revealed a high positive correlation of deaths with percentage of African Americans and unclear relationship with racial majority which i primarily Caucasian in the United States(Figure 1 & 2 a, b). Direct flights from China, especially Wuhan, were also significant in both models as predictors of death, therefore reflecting early spread of the disease(Figure 1 & 2 a, b).

**Figure 2a.**
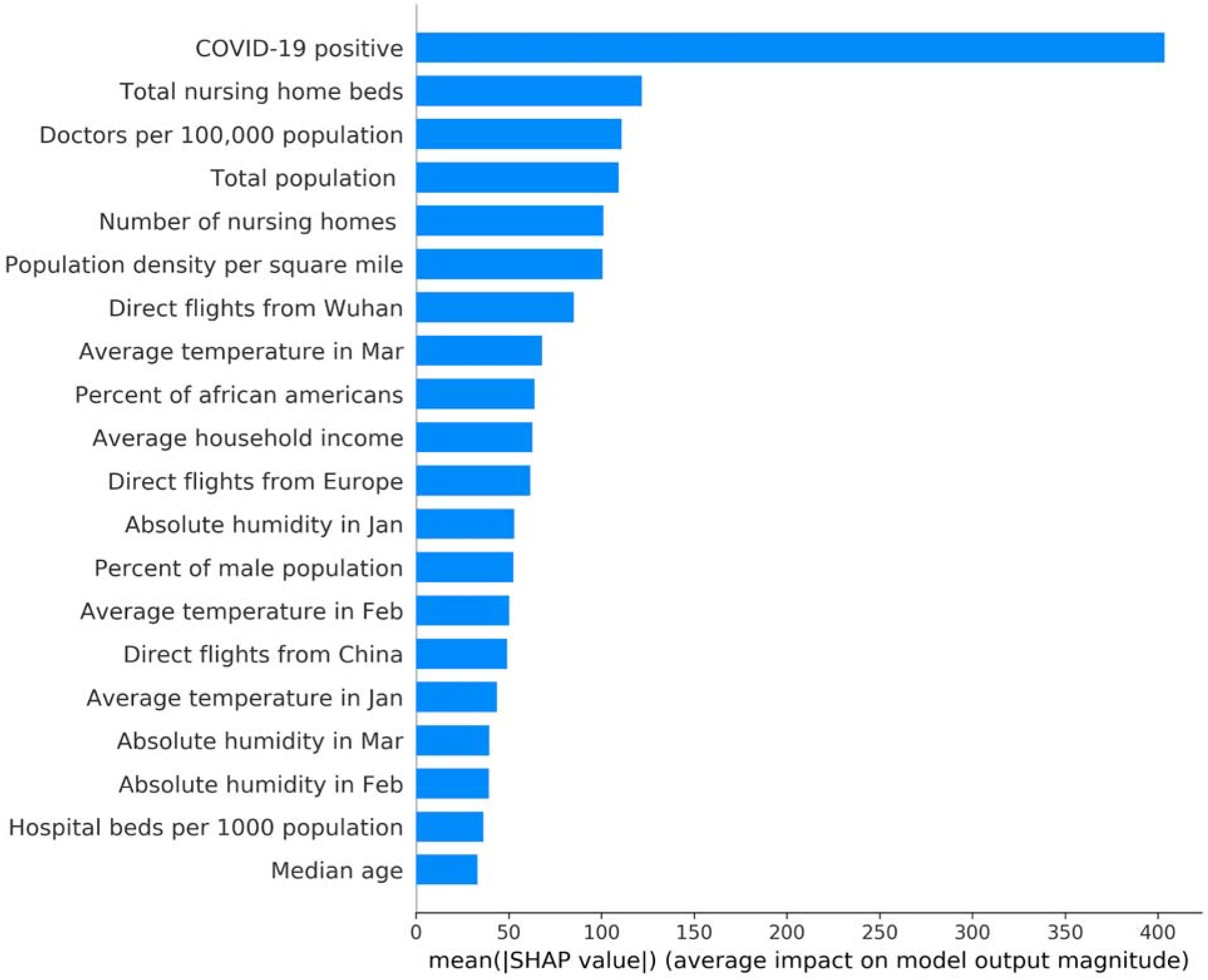
SHAP feature importance model results for COVID-19 deaths in the US. Feature importance plot lists the most significant features in descending order. The top features contribute more to the model than the bottom ones and thus have high predictive power.^11^

**Figure 2b.**
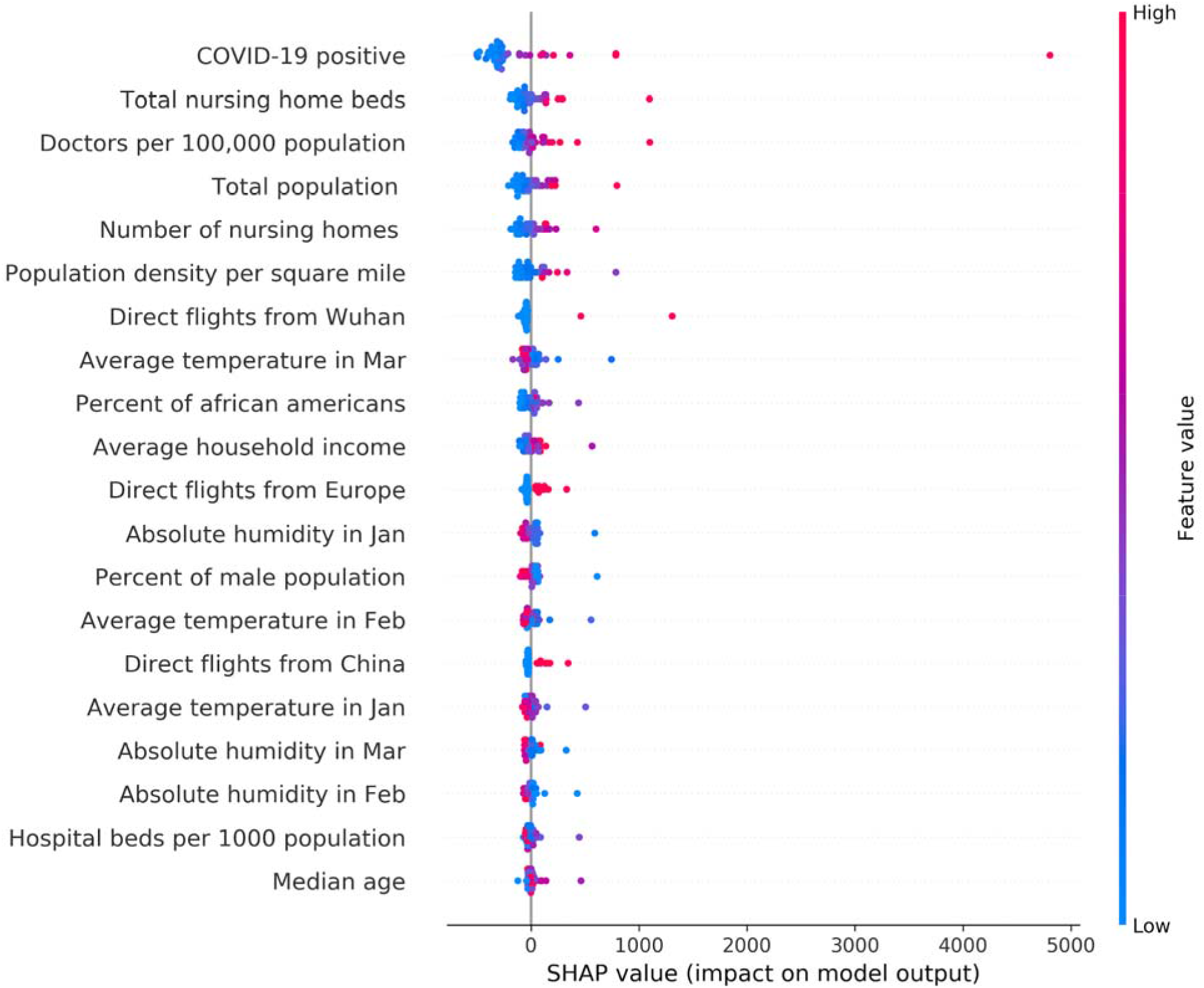
SHAP feature importance distribution amongst each state for COVID-19 deaths in the US. The vertical location of the dots show the feature they are depicting with each dot representing a state value.Color shows whether that feature value was high(red) or low(blue) for that row of the dataset.Horizontal location shows whether the effect of that value caused a higher or lower prediction.^11^

Associations between deaths and weather patterns, hospital bed capacity, median age, timing of administrative action to mitigate disease spread such as the closure of educational institutions or stay at home order were not significant.

## Discussion

COVID-19 disease has varied spread and mortality across communities amongst different states in the US. While multiple mortality prediction models with variable accuracy are available, our goal is to identify state specific key determinants of COVID-19 mortality to guide disease mitigation and management efforts.^13^ Our approach is transparent and explainable by providing relative weight of each feature in predicting mortality, thereby helping local and national policymakers manage this pandemic effectively.

The strength of this study is implementation of two independently developed machine learning models and separate feature importance algorithms resulting in similar results validating our findings. Also, our findings have face validity based on real life observations of actual mortality patterns across the states and within certain geographical locations such as New York City.^14,15^ Our models show that high population density, number of nursing home beds and foreign travel may increase transmission and thus COVID-19 mortality. Transmission and spread of the SARS-CoV-2 virus has followed the three key features since identification of COVID-19 disease in the United States. Results of analysis also demonstrate the high importance of direct flights from China, specifically from the Wuhan province. Despite the travel ban in China and to the US, this travel pattern had marked effect on the international scale.^16^ With the European strain of the virus associated with New York’s disease transmission, variations in the relative feature importance of the two machine learning models amongst direct flights from China and Europe, might also reflect the variable pattern of transcontinental spread of the disease to the US.^17^ Once, SARS-CoV-2 virus arrived in the US, it clearly impacted areas of high population density across the country as reported by the CDC.^2^ Within these high population density locations, clusters of nursing home and their residents have been significantly impacted with much higher rates of mortality.^18^ Along with clear association with high number of nursing home patients, doctors per 100,000 population also reflects upon the possibility of higher number of population with significant comorbidities requiring medical management. It is unclear why hospital beds per capita had low feature importance in the results, although we would have expected the same association as doctors per capita.Further analysis using more healthcare data might explain this better. Understanding such patterns using machine learning techniques can empower public health officials to deploy targeted restrictions.

Similarly, percentage of African American amongst the population was also found to be of high importance in prediction of COVID-19 mortality, as described in various other analyses.^14,15^ This could be due to many reasons and may have many confounders such as median income, higher rate of pre-existing disease and higher populations in areas with higher population density areas.^15^ Improved access to care in the near term and addressing concerns surrounding social determinants of health in the medium and longer term is likely to address these long standing disparities and related higher mortality.The effect of other features such as median age, climate and mitigation measures on COVID-19 related mortality is not clear based on our analysis.

The effect of other features such as median age,climate and mitigation measures on COVID-19 related mortality is not clear based on our analysis.For example, the low feature importance of administrative actions to mitigate disease spread and its resulting mortality is surprising.But in our study,we may not have seen the impact of mitigation measures since the mortality data for this study was limited till April 30,2020 and most of these measures went into effect in the months of March and April across the US.and may reflect delayed outcomes of interventions which were not yet reflected in current data used in our models. Median age is consistent across states thus low importance is understandable, also most COVID-19 related mortalities in the US have occurred in elderly population.^19^ Distribution of weather pattern in the US during the time frame of this study would have had limited impact, if any, on SARS-CoV-2 virus transmission supporting our finding of limited effects on COVID-19 mortality.^20^

Beyond prediction of disease related outcomes, machine learning models are being increasingly utilized with explainable and interpretable results in healthcare.^5,21,22^ Use of multiple models and comparison of results has become more common in healthcare as investigators seek to overcome concerns around generalizability and accuracy of the results.^21,23^ Although random forest has been shown to handle even imbalanced data, newer machine learning models such as CatBoost in addition have also demonstrated capability of handling different types of variables while maintaining high level of accuracy and efficiency.^8,24^ Explainable and interpretable results from machine learning models may not demonstrate causality but provide better insight into the results of machine learning models, overcoming some of the concerns around machine learning being a black box.^25^ Feature engineering algorithms such as SHAP and Boruta algorithm are being increasingly utilized in various fields of healthcare to ascertain feature importance and deliver personalized prediction models.^5^

## Conclusion

COVID-19 disease has heterogeneous clinical presentation and variable spread across different locations. Understanding the determinants of COVID-19 outcomes, using dynamic, scalable and explainable machine learning models can help guide resource management and policy framework.^3^

## Data Availability

Data sources listed in supplemental content

## Notes

### Competing Interest Statement

The authors have declared no competing interest.

### Funding Statement

No external funding was received.

### Author Declarations

IRB approval not required.

## References

1. Schuchat A, Team CC-R. Public Health Response to the Initiation and Spread of Pandemic COVID-19 in the United States, February 24-April 21, 2020. MMWR Morb Mortal Wkly Rep. 2020;69(18):551–556.

2. Team CC-R. Geographic Differences in COVID-19 Cases, Deaths, and Incidence - United States, February 12-April 7, 2020. MMWR Morb Mortal Wkly Rep. 2020;69(15):465–471.

3. Bedford J, Enria D, Giesecke J, et al. COVID-19: towards controlling of a pandemic. Lancet (London, England). 2020;395(10229):1015–1018.

4. Sanders JM, Monogue ML, Jodlowski TZ, Cutrell JB. Pharmacologic Treatments for Coronavirus Disease 2019 (COVID-19): A Review. JAMA. 2020.

5. Hilton CB, Milinovich A, Felix C, et al. Personalized predictions of patient outcomes during and after hospitalization using artificial intelligence. NPJ Digit Med. 2020;3:51.

6. Gupta S, Raghuwanshi GS, Chanda A. Effect of weather on COVID-19 spread in the US: A prediction model for India in 2020. The Science of the total environment. 2020;728:138860.

7. Breiman L. Random forests. Machine learning. 2001;45(1):5–32.

8. Prokhorenkova L, Gusev G, Vorobev A, Dorogush AV, Gulin A. CatBoost: unbiased boosting with categorical features. Proceedings of the 32nd International Conference on Neural Information Processing Systems; 2018; Montréal, Canada.

9. Hamilton DF, Ghert M, Simpson AH. Interpreting regression models in clinical outcome studies. Bone Joint Res. 2015;4(9):152–153.

10. Kursa MB, Rudnicki WR. Feature Selection with the Boruta Package. 2010. 2010;36(11):13.

11. Lundberg SM, Lee S-I. A unified approach to interpreting model predictions. Proceedings of the 31st International Conference on Neural Information Processing Systems; 2017; Long Beach, California, USA.

12. Li H, Wang S, Zhong F, et al. Age-Dependent Risks of Incidence and Mortality of COVID-19 in Hubei Province and Other Parts of China. Front Med (Lausanne). 2020;7:190.

13. Jewell NP, Lewnard JA, Jewell BL. Caution Warranted: Using the Institute for Health Metrics and Evaluation Model for Predicting the Course of the COVID-19 Pandemic. Annals of Internal Medicine. 2020.

14. Wadhera RK, Wadhera P, Gaba P, et al. Variation in COVID-19 Hospitalizations and Deaths Across New York City Boroughs. JAMA. 2020.

15. Yancy CW. COVID-19 and African Americans. JAMA. 2020.

16. Chinazzi M, Davis JT, Ajelli M, et al. The effect of travel restrictions on the spread of the 2019 novel coronavirus (COVID-19) outbreak. Science. 2020;368(6489):395–400.

17. Gonzalez-Reiche AS, Hernandez MM, Sullivan MJ, et al. Introductions and early spread of SARS-CoV-2 in the New York City area. Science. 2020.

18. Gandhi M, Yokoe DS, Havlir DV. Asymptomatic Transmission, the Achilles’ Heel of Current Strategies to Control Covid-19. The New England journal of medicine. 2020;382(22):2158–2160.

19. Promislow DEL. A geroscience perspective on COVID-19 mortality. J Gerontol A Biol Sci Med Sci. 2020.

20. Wu Y, Jing W, Liu J, et al. Effects of temperature and humidity on the daily new cases and new deaths of COVID-19 in 166 countries. The Science of the total environment. 2020;729:139051.

21. Mathur P, Burns ML. Artificial Intelligence in Critical Care. Int Anesthesiol Clin. 2019;57(2):89–102.

22. Ahmed Z, Mohamed K, Zeeshan S, Dong X. Artificial intelligence with multi-functional machine learning platform development for better healthcare and precision medicine. Database : the journal of biological databases and curation. 2020;2020.

23. Wang Y, Lei L, Ji M, Tong J, Zhou CM, Yang JJ. Predicting postoperative delirium after microvascular decompression surgery with machine learning. J Clin Anesth. 2020;66:109896.

24. Khalilia M, Chakraborty S, Popescu M. Predicting disease risks from highly imbalanced data using random forest. BMC Med Inform Decis Mak. 2011;11:51.

25. Vellido A. Societal Issues Concerning the Application of Artificial Intelligence in Medicine. Kidney Dis (Basel). 2019;5(1):11–17.

